# Electronic Nicotine Delivery Systems (ENDS) use during a five-year period is not associated with self-reported chronic obstructive pulmonary disease (COPD) after adjustment of cigarette smoking history: A longitudinal analysis of PATH data

**DOI:** 10.1101/2022.02.20.22271250

**Authors:** Steven F. Cook, Jana L. Hirschtick, Nancy L. Fleischer, Douglas A. Arenberg, Geoffrey D. Barnes, David T. Levy, Luz Maria Sanchez-Romero, Jihyoun Jeon, Rafael Meza

**Affiliations:** Department of Epidemiology, University of Michigan, Ann Arbor, MI; Division of Pulmonary and Critical Medicine, Department of Internal Medicine, University of Michigan, Ann Arbor; Department of Internal Medicine, Frankel Cardiovascular Center, University of Michigan Health System, Ann Arbor; Institute for Healthcare Policy and Innovation, University of Michigan, Ann Arbor; Department of Oncology, Georgetown University, Washington, DC

**Author notes:** **Corresponding author**:; mail: Department of Epidemiology, 1415 Washington Heights, Ann Arbor, MI, 48109, United States.

**Keywords:** ENDS, cigarettes, COPD, respiratory disease

## Abstract

**Background:** Understanding the relationship between electronic nicotine delivery systems (ENDS) use and chronic obstructive pulmonary disease (COPD) and other respiratory conditions is critical. However, previous studies have not adequately controlled for history of cigarette smoking.

**Research Question:** To examine the prospective association between ENDS use and self-reported incident COPD after adjusting for cigarette smoking history.

**Study Design and Methods:** Using waves 1-5 of the US Population Assessment of Tobacco and Health (PATH) study, we examined the association between ENDS use and self-reported incident COPD among adults aged 40+ using discrete time survival models. Current ENDS use was measured as a time-varying covariate, lagged by one wave, defined as established daily or some days use. We controlled for baseline demographics (age, sex, race/ethnicity, education), health characteristics (asthma, obesity, exposure to second-hand smoke), and smoking history (smoking status and cigarette pack-years).

**Results:** Incident COPD was self-reported by 925 respondents during the five-year follow-up period. Prior to adjusting for other covariates, time-varying ENDS use appeared to nearly double the risk of incident COPD (HR 1.98, 95% CI 1.44-2.74). However, ENDS use was no longer significantly associated with COPD (aHR 1.10, 95% CI 0.78-1.57) after adjusting for current cigarette smoking and cigarette pack-years. The risk of self-reported incident COPD increased with cigarette pack-years and was higher for respondents who were older, female, less educated, and had baseline asthma or obesity.

**Interpretation:** ENDS use did not significantly increase the risk of self-reported incident COPD over a five-year period once current smoking status and cigarette pack-years were taken into account. Cigarette pack-years, on the other hand, remained associated with a net increase in the risk of self-reported incident COPD. These findings highlight the importance of using prospective longitudinal data and properly controlling for cigarette smoking history to assess the independent health effects of ENDS.

## Introduction

Chronic Obstructive Pulmonary Disease (COPD) is a chronic and progressive respiratory disease that encompasses the clinical definitions of emphysema and chronic bronchitis.^1^ COPD is characterized by expiratory airflow limitation and abnormal airway inflammation,^2^ usually caused by exposure to noxious particles or gases.^3^ COPD is the fourth leading cause of mortality in the United States,^4^ and is projected to be responsible for 9.42 million deaths and more than $800 billion in direct medical costs by 2038.^5^

Cigarette smoking is the primary preventable risk factor for COPD,^3,6^ with more than 20% of long-term tobacco users expected to develop COPD.^2,6^ The risk of COPD is 200% and 30% higher in current smokers than in non-smokers and former smokers, respectively,^7^ and the risk of developing COPD has been also associated with second-hand smoke (SHS) exposure.^8^ Years of chronic smoking are needed to develop COPD,^6,9^ with recent research demonstrating that smoking duration and intensity are both important determinants of COPD risk.^10^ Years since quit, on the other hand, is more weakly associated with COPD risk,^10^ suggesting that the damage caused by long-term smoking is cumulative and the impact may be hard to reverse.

Electronic Nicotine Delivery Systems (ENDS) product use has been widespread in the US since about 2012,^11^ with growing concern that ENDS use may also increase the risk of COPD.^12^ However, most epidemiologic research on the ENDS-COPD association has been based on cross-sectional studies,^13–16^ which are limited by potential reverse causation.^17^ Without information on the timing of both exposure and outcome, it is not possible to know whether ENDS use came before or after COPD diagnosis. In particular, adults who smoke might switch to ENDS after being diagnosed with COPD.^18–20^ Moreover, most adults who use ENDS are either currently smoking cigarettes or have smoked cigarettes in the past,^21,22^ making it important to properly account for their risk due to current or past cigarette smoking when assessing any potential independent risks from ENDS use. Therefore, cross-sectional studies need to be interpreted with caution, and prospective longitudinal data with detailed cigarette smoking histories, including duration, intensity, and time since quit, are needed to better understand any potential independent effects of ENDS use on COPD risk.

In this study, we analyze data from five waves of a US nationally representative longitudinal cohort study to examine the prospective association between ENDS use and self-reported diagnosed incident COPD among adults aged 40 and older. Our study: (1) examines the incidence of COPD prospectively, limiting concerns with reverse causation; (2) includes ENDS use as a one-wave-lagged time-varying measure to ensure that ENDS use preceded the COPD diagnosis; and (3) controls for the duration, intensity, and history of cigarette smoking.

## Methods

### Data

We used data on adults from Waves 1-5 (2013-2019) of the Population Assessment of Tobacco and Health (PATH) Study, a nationally representative longitudinal study of the non-institutionalized civilian US population. Wave 1 data were collected from September, 2013 to December, 2014; Wave 2 data were collected from October, 2014 to October, 2015; Wave 3 data were collected from October, 2015 to October, 2016; Wave 4 data were collected from December, 2016 to January, 2018; and Wave 5 data were collected from December, 2018 to November 2019. Analyses were conducted using the restricted-access PATH adult data files.^23^ Further details on the survey design, including how to access restricted use data, are described elsewhere.^24,25^

We analyzed self-reported diagnosed COPD incidence over four waves of follow-up (Wave 2-Wave 5). Since COPD is rare in young adults,^26^ PATH suppressed COPD outcomes for respondents under age 40 beginning in Wave 4. We therefore restricted the analytic sample to respondents aged 40 or older who reported no history of any COPD outcome (i.e., COPD, chronic bronchitis, and emphysema) at baseline (Wave 1) and who completed at least one follow-up interview. Respondents were censored at the time of their first self-reported COPD outcome at follow-up. Those who did not report an outcome were censored at their last observation point. The final analytic sample consisted of 9,861 respondents. A flowchart summarizing the analytic sample is provided in the supplemental material (Figure S1).

### Self-Reported Diagnosed Incident COPD

We examined self-reported diagnosed COPD incidence at each follow-up interview based on the question: “*In the past 12 months, has a doctor, nurse, or other health professional told you that you had…(1) COPD, (2) chronic bronchitis, or (3) emphysema*?” Consistent with the clinical definition of COPD, respondents who reported having any of these conditions were considered to have COPD.

In Waves 2 and 3, *<u>all</u>* respondents were asked, *“In the past 12 months, has a doctor, nurse or other health professional told you that you had…COPD, chronic bronchitis, or emphysema?”*. Due to a change in the skip pattern in Waves 4 and 5, this question was *<u>only</u>* asked to respondents who reported seeing a *“medical doctor, nurse, or other health professional”* during the past 12 months. For Waves 2 and 3, we adopted an inclusive measurement strategy, classifying all respondents who reported a COPD outcome regardless of whether they saw a health professional in the past 12 months. In Waves 4 and 5, we classified respondents who did not see a health professional in the past 12 months, and were therefore not asked about COPD, as not having the outcome. As a sensitivity analysis, we also restricted the COPD outcome to respondents who self-reported COPD and reported seeing a doctor during the past 12 months.

### Electronic nicotine delivery systems (ENDS)

The ENDS use exposure variable was based on self-reported every day or someday use among established users (ever fairly regularly used ENDS). This variable was measured at each wave and included as a time-varying exposure. To ensure that ENDS use preceded the COPD outcome, we lagged the ENDS exposure variable by one wave (t-1).

### Smoking variables

To account for the potential confounding effect of cigarette smoking, two smoking history covariates were included. First, using established cigarette use (100 or more cigarettes smoked in lifetime), we created a smoking status variable (non-smokers, former smokers, current smokers). Non-smokers included non-established smokers and never smokers. Former smokers were defined as established smokers who did not currently smoke. Current smokers were defined as established smokers who currently smoked cigarettes every day or some days. To capture changes in cigarette smoking over time, the smoking status variable was included as a time-varying covariate. Second, we included cigarette pack-years (CPY) at Wave 1 as a measure of lifetime cigarette smoking at baseline. CPY was calculated by multiplying the reported years of cigarette smoking by the average number of packs-per-day while individuals smoked. CPY for non-smokers were coded as 0. Respondents who reported smoking more than 200 cigarettes per day (10 packs per day) were considered implausible and were set to missing. Preliminary analyses suggested that a log transformation of the CPY measure best fit the data, which is the functional form we employed in the final models.

### Other Covariates

We included age (continuous), sex (male, female), race/ethnicity (Hispanic, Non-Hispanic (NH) White, NH Black, NH Other), education (high school or less, some college, bachelor’s degree/graduate degree), and health insurance status (some vs. none) as baseline sociodemographic covariates. We also included obesity (Body Mass Index (BMI) >=30 vs. < 30) and asthma as baseline COPD risk factors. To capture variation in exposure to second-hand smoke, we included the number of hours respondents reported ‘close’ exposure to second-hand smoke during the past 7 days. Respondents who reported ‘don’t know’ were considered to have no close exposure, and the number of hours exposed during the past 7 days were top coded at 100 (range 0-100). Second-hand smoke exposure was included as a time-varying covariate.

### Statistical Analysis

Descriptive statistics were first calculated for sociodemographic characteristics, smoking behaviors and COPD risk factors at baseline. Sample characteristics were then calculated according to respondent’s cigarette/ENDS use at baseline. Next, lifetables were used to describe the distribution of self-reported diagnosed incident COPD at follow-up (Waves 2-5). The discrete-time baseline hazard estimates reflect the weighted conditional probability of COPD in the risk set at each discrete time period.^27^

We fitted a series of multivariable discrete-time survival models predicting self-reported incident COPD at follow-up (Wave 2 to Wave 5). The 9,861 respondents in our analytic sample were restructured to an unbalanced person-period data set where each respondent *(N)* contributed a separate row of data for each discrete-time interval (*T*), with a maximum of four rows per respondent, until they were diagnosed with COPD or right censored. The person period dataset, constructed based on *N x T*, had 33,679 observations, and provided the data structure for the discrete-time survival analyses. Four discrete-time survival models were estimated using a complimentary log-log link function on the person-period data. Model 1 included the unadjusted ENDS use exposure variable. Model 2 added the sociodemographic control variables, COPD risk factors and time-varying second-hand smoke exposure. Model 3 added time-varying smoking status, and Model 4 added cigarette pack-years.

Data were weighted using Wave 1 weights, including full-sample and 100 replicate weights, to ensure that results were representative of the non-institutionalized US adult population at baseline. To assess the impact of attrition, we compared baseline characteristics for respondents with complete and incomplete follow-up data (Appendix, Table A1). Because respondents with incomplete follow-up data had a slightly different sociodemographic profile than respondents who participated at each wave of follow-up, we also fitted discrete time models using the ‘all waves weights’ as a sensitivity analysis. These weights account for this type of attrition^24^ and restricts the analysis to a longitudinal cohort of respondents who participated in all waves of the PATH study (person n=7065, risk period n=27,190). Additionally, we conducted another sensitivity analysis that restricted the COPD outcome to respondents who also reported seeing a doctor during the past 12 months. For all analyses, variances were computed using the balanced repeated replication methods with Fay’s adjustment set to 0.3 as recommended by the PATH study.^28,29^ All analyses were conducted using Stata 17.1.^30^

## Results

The baseline sociodemographic characteristics, COPD risk factors and tobacco variables for the analytic sample (n=9,861) are outlined in Table 1. At baseline (Wave 1, 2013-14), respondents had a mean age of 57.4 years (SD=11.9), 53% were female, 70.4% were NH White, 11.2% were NH Black, and 11.4% were Hispanic. Regarding education, 60.6% had at least some college education, while 39.4% reported completing a high school education or less. Approximately one-third of respondents had a BMI of 30 kg/m^2^ or more (33.5%), while 8.7% reported a lifetime diagnosis of asthma at baseline. Every day or someday use of ENDS was reported by 1.4% of respondents at baseline. Most respondents were never established cigarette smokers at baseline (63.0%), while 23.2% were former and 13.8% were current cigarette smokers. Among current or former established smokers, the average cigarette pack-years at baseline was 23.9 (SD=26.4) In terms of exposure to second-hand smoke, 41% reported past 7-day exposure to second-hand smoke, with an average 10.1 hours (SD=22.2) of exposure.

**Table 1.**
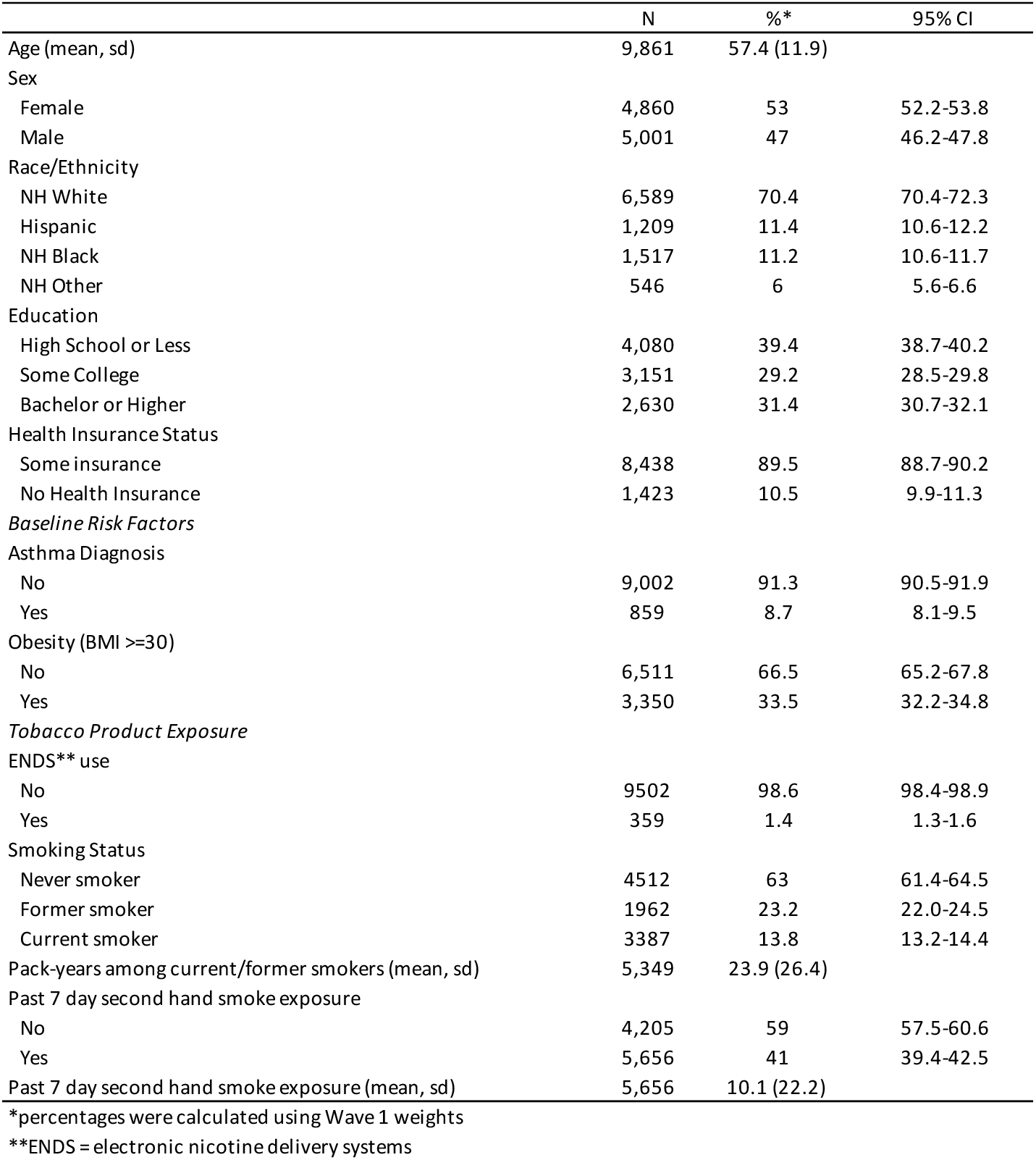
Weighted sociodemographic characteristics, COPD risk factors, and smoking behaviors for adult respondents (40+), Population Assessment of Tobacco &Health Study (Wave 1, 2013-2014)

Table 2 presents lifetables describing the incidence of self-reported COPD, reflecting the conditional probability of COPD diagnosis at each discrete time interval. In total there were 925 self-reported incident COPD cases, with an average annualized incidence of 1.97% (range 1.4%-2.4%) during the five-year follow-up period.

**Table 2.** Life table describing the incidence of self-reported COPD among adults (40+), Population Assessment of Tobacco and Health Study (Waves 1-5, 2013-2019)

| Interval | Total | COPD Diagnosis | Censored | Survivor Estimate* | Hazard Estimate** |
| --- | --- | --- | --- | --- | --- |
| Period 1 (W1-W2) | 9,861 | 314 | 646 | 0.968 | 0.024 |
| Period 2 (W2-W3) | 8,901 | 252 | 719 | 0.941 | 0.021 |
| Period 3 (W3-W4) | 7,930 | 158 | 785 | 0.922 | 0.014 |
| Period 4 (W4-W5) | 6,987 | 201 | 6,785 | 0.896 | 0.019 |

Notes:
\*survival estimates were based on unweighted data
\*\*hazard estimates were calculated with the replicate weights

Table 3 presents the results of the four models examining the risk of self-reported incident COPD across the five-year follow-up period. Prior to adjusting for other covariates (Model 1), time-varying ENDS use appeared to nearly double the risk of incident COPD (HR 1.98, 95% CI 1.44-2.74). However, ENDS use was no longer significantly associated with COPD risk (aHR 1.17, 95% CI 0.83-1.66) after adjusting for current cigarette smoking in Model 3 and additionally adjusting for cigarette pack-years in Model 4 (aHR 1.10, 95% CI 0.78-1.57).

**Table 3.** Discrete time survival analysis predicting incidence of self-reported chronic obstructive pulmonary disease (COPD) for respondents aged 40 and older, Population Assessment of Tobacco and Health Study (Waves 1-5, 2013-2019)

|  | Model 1<sup>a</sup> |  | Model 2<sup>b</sup> |  | Model 3<sup>c</sup> |  | Model 4<sup>d</sup> |  |
| --- | --- | --- | --- | --- | --- | --- | --- | --- |
|  | Hazard | 95% CI | Hazard | 95% CI | Hazard | 95% CI | Hazard | 95% CI |
| *Time-varying ENDS use* | 1.98*** | 1.44-2.74 | 1.70** | 1.21-2.40 | 1.17 | .83-1.66 | 1.1 | .78-1.57 |
| Age† |  |  | 1.03*** | 1.02-1.03 | 1.03*** | 1.02-1.04 | 1.03*** | 1.02-1.04 |
| Sex (Female=1) |  |  | 1.59*** | 1.34-1.88 | 1.68*** | 1.41-2.00 | 1.79*** | 1.50-2.13 |
| Race/Ethnicity |  |  |  |  |  |  |  |  |
| NH White |  |  | REF | REF | REF | REF | REF | REF |
| Hispanic |  |  | 0.94 | .73-1.21 | 1.05 | .82-1.36 | 1.18 | .92-1.52 |
| NH Black |  |  | 1.21 | .99-1.49 | 1.17 | .94-1.44 | 1.27* | 1.03-1.58 |
| NH Other |  |  | 0.94 | .59-1.49 | 0.99 | .63-1.56 | 1 | .64-1.59 |
| Education |  |  |  |  |  |  |  |  |
| High School or Less |  |  | 2.19*** | 1.67-2.88 | 1.83*** | 1.40-2.40 | 1.72*** | 1.31-2.26 |
| Some College |  |  | 1.57** | 1.17-2.10 | 1.40* | 1.05-1.88 | 1.33* | 1.0-1.78 |
| Bachelor Degree or Higher |  |  | REF | REF | REF | REF | REF | REF |
| Uninsured |  |  | 1.32 | .99-1.76 | 1.18 | .88-1.57 | 1.16 | .88-1.55 |
| *Baseline Risk Factors* |  |  |  |  |  |  |  |  |
| Asthma |  |  | 2.95*** | 2.26-3.84 | 2.99*** | 2.29-3.90 | 3.05*** | 2.33-4.00 |
| Obesity (BMI>30) |  |  | 1.34** | 1.11-1.63 | 1.43*** | 1.18-1.73 | 1.41** | 1.16-1.71 |
| Time-varying second-hand smoke exposure†† |  |  | 1.19*** | 1.15-1.23 | 1.11*** | 1.07-1.16 | 1.09*** | 1.04-1.14 |
| *Time-Varying Smoking Status Indicator* |  |  |  |  |  |  |  |  |
| Never smoker |  |  |  |  | REF | REF | REF | REF |
| Former Smoker |  |  |  |  | 1.50** | 1.17-1.94 | 0.85 | .59-1.23 |
| Current Smoker |  |  |  |  | 3.16*** | 2.58-3.87 | 1.64** | 1.17-2.29 |
| Log cigarette pack-years †† |  |  |  |  |  |  | 1.79*** | 1.46-2.19 |

Notes: Person N=9,861; risk N=33,679; \*p<0.05, \*\*p<0.01, \*\*\*p<0.001
<sup>a</sup> ENDS exposure <sup>b</sup> sociodemographic variables added; <sup>c</sup> smoking status added; <sup>d</sup> cigarette pack-years added
† age mean centered for models
†† variable rescaled to reflect intervals of 10

Multivariable associations between other variables and COPD can also be found in Model 4, Table 3. Self-reported incident COPD increased with the log of cigarette pack-years (aHR 1.79, 95% CI 1.46-2.19) and was higher for respondents who were older (aHR 1.03, 95% CI 1.02-1.04), female (aHR 1.79, 95% CI 1.50-2.13), had a high school degree or less (aHR 1.72, 95% CI 1.31-2.26), and had baseline asthma (aHR 3.05, 95% CI 2.33-4.00) or obesity (aHR 1.41, 95% CI 1.16-1.71). Exposure to second-hand smoke was also associated with the risk of incident COPD, as every 10-hours of exposure increased the risk of COPD by 9.0% (aHR 1.09, 95% CI 1.04-1.14). Current smoking status remained significant after adjusting for the log of cigarette pack-years in Model 4 (aHR 1.64, 95% CI 1.17-2.29).

The confounding effect of smoking on the association between ENDS use and self-reported incident COPD can be further seen in Table 4. Most adults who used ENDS at each wave were either current (range 49.7%-61.4%, decreasing by Wave) or former (range 30.7%-46.7%, increasing by Wave) established cigarette smokers, while less than 8% were never established smokers (range 3.4%-7.9%, decreasing by Wave). Most adults who did not use ENDS, on the other hand, were never established cigarette smokers (range 60.5%-63.8%). Moreover, most adults who used ENDS had significantly higher baseline cigarette pack-years and higher levels of second-hand smoke exposure than adults who did not use ENDS.

**Table 4.** Time-varying ENDS use by smoking status, cigarette pack-years and second-hand smoke exposure, Population Assessment of Tobacco and Health Study (Waves 1-5, 2013-2019)

|  | Wave 1 |  |  |  |  | Wave 2 |  |  |  |  | Wave 3 |  |  |  |  | Wave 4 |  |  |  |  |
| --- | --- | --- | --- | --- | --- | --- | --- | --- | --- | --- | --- | --- | --- | --- | --- | --- | --- | --- | --- | --- |
|  | No ENDS use |  | ENDS use |  | P | No ENDS use |  | ENDS use |  | P | No ENDS use |  | ENDS use |  | P | No ENDS use |  | ENDS use |  | P |
|  | % or mean | 95% CI | % or mean | 95% CI |  | % or mean | 95% CI | % or mean | 95% CI |  | % or mean | 95% CI | % or mean | 95% CI |  | % or mean | 95% CI | % or mean | 95% CI |  |
| Prevalence of ENDS use | 98.6 | 98.4-98.7 | 1.4 | 1.2-1.6 |  | 98.3 | 98.0-98.5 | 1.7 | 1.5-2.0 |  | 98.3 | 98.1-98.5 | 1.7 | 1.5-1.9 |  | 98.6 | 98.3-98.8 | 1.4 | 1.2-1.7 |  |
| *Time-Varying Smoking Status Indicator* |  |  |  |  | *** |  |  |  |  | *** |  |  |  |  | *** |  |  |  |  | *** |
| Proportion Never smoker (%) | 63.8 | 62.2-65.3 | 7.9 | 5.5-11.1 |  | 61.6 | 60.0-63.1 | 4.1 | 2.4-7.1 |  | 60.7 | 59.0-62.3 | 3.4 | 1.8-6.2 |  | 60.5 | 58.7-62.3 | 3.6 | 1.6-7.8 |  |
| Proportion Former Smoker (%) | 23.1 | 21.9-24.4 | 30.7 | 25.3-36.7 |  | 25.2 | 23.9-26.6 | 37.5 | 31.8-43.7 |  | 26.3 | 24.9-27.8 | 40.8 | 34.7-47.2 |  | 27.2 | 25.6-28.8 | 46.7 | 39.6-54.0 |  |
| Proportion Current Smoker (%) | 13.1 | 12.5-13.7 | 61.4 | 55.9-66.7 |  | 13.2 | 12.6-13.8 | 58.4 | 52.6-63.9 |  | 13 | 12.3-13.6 | 55.8 | 49.7-61.8 |  | 12.3 | 11.6-13.1 | 49.7 | 42.4-57.1 |  |
| Baseline cigarette pack-years (mean, sd) | 24.5 (26.1) |  | 27.4 (32.4) |  | * | 23.7 (25.5) |  | 29.2 (32.7) |  | ** | 22.9 (24.7) |  | 28.5 (32.9) |  | ** | 22.3 (24.1) |  | 30.9 (35.5) |  | ** |
| Time-varying second-hand smoke exposure (mean, sd) | 4.0 (12.7) |  | 12.3 (34.4) |  | *** | 3.5 (11.8) |  | 13.1 (35.7) |  | *** | 3.1 (10.8) |  | 11.2 (31.4) |  | *** | 2.9 (10.3) |  | 9.9 (29.6) |  | *** |

\*p<0.05, \*\*p<0.01, \*\*\*p<0.001

### Sensitivity Analyses

The discrete-time models were estimated using the longitudinal cohort with the ‘all waves’ weights, which resulted in a reduced sample size because participation was required in all five waves (see Table S1). The substantive results were nearly identical as effect of ENDS use was no longer significant after adjusting for current cigarette smoking status and for cigarette pack-years. As a secondary sensitivity analysis, discrete-time models were estimated with the COPD outcome restricted to those who also reported seeing a health care professional during the past 12 months in Waves 2 and 3, consistent with the definition in Waves 4 and 5 (see Table S2). The substantive results were again nearly identical, as ENDS use was not associated with incident COPD in the fully adjusted model. Finally, we defined e-cigarette use as 10+ days in the past 30 days as a sensitivity analysis rather than every day or someday use. The results using this ENDS use definition were similar (see Table S3).

## Discussion

This study examined the prospective association between ENDS use and self-reported incident COPD over a five-year follow-up period. Prior to adjusting for current and historical cigarette use, ENDS use appeared to be associated with an increased risk of incident COPD in US adults. This unadjusted estimate is consistent with findings from existing cross-sectional research.^13–16^ While previous studies typically included some measure of current smoking status, none controlled for smoking duration or intensity. High levels of smoking exposure are needed for COPD to develop,^10,31,32^ and not controlling for this exposure may potentially conflate the effect of ENDS use with individual smoking histories. In our study, once we adjusted for time-varying smoking status and baseline smoking history, measured as pack-years, ENDS use was no longer associated with incident COPD. This finding is not surprising since more than 90% of adults who used ENDS aged 40 or over in our sample were either current or former established cigarette smokers at each discrete time interval. Moreover, the average pack-years of adults who used ENDS who were established smokers was significantly higher than the average pack-years values for adults who were current and former smokers and who did not use ENDS. These findings, considered together, demonstrate the importance of properly accounting for both smoking status and smoking histories when studying the health effects of ENDS.

The confounding effect of cigarette smoking histories in the ENDS-COPD association is compounded by the inability of cross-sectional studies to account for the relative timing of ENDS use and COPD outcomes. This limitation is endemic to cross-sectional research of health effects, but it is particularly important in research on the chronic respiratory health effects of ENDS use because some long-term smokers might have switched to ENDS products after experiencing adverse health outcomes, including COPD.^19,20,33^ By restricting our analytic sample to respondents without COPD at baseline and examining incident COPD prospectively, our study was able to limit reverse causation concerns. In addition, because smokers may be diagnosed with COPD and switch to ENDS products in the same year, we lagged our ENDS exposure by one wave to ensure that ENDS use preceded the COPD outcome.

Consistent with literature,^6,7,34^ we found a strong association between smoking status and COPD, with higher risk among adults who currently smoked than adults who smoked in the past or never smoked.^7^ However, smoking status by itself is limited because it does not account for the level of exposure. We know that long-term smoking is required to develop COPD,^9^ and our findings confirm that cigarette pack-years is independently associated with the risk of incident COPD. Similar to other research,^8^ we also found that time-varying second-hand smoke exposure was predictive of self-reported diagnosed incident COPD. It has been argued that more detailed information on smoking histories are needed to improve the predictive accuracy of COPD risk models,^10^ and incorporating measures of both direct and indirect smoking exposure are important to more fully understand the smoking-COPD relationship.

This study is among the first to examine prospective associations between ENDS use and COPD while formally accounting for current and historical cigarette use. Despite this contribution, there are several limitations to note. First, the findings were based on approximately five years of data and a longer follow-up may be required to fully understand the role of ENDS use on the risk of developing COPD, a chronic and long-term condition. If similar exposure time is required for ENDS as is required for cigarettes, it is possible that the downstream consequences of ENDS use may not be observable until far into the future.^9^ Not only are ENDS products relatively new to the tobacco marketplace, they also continue to evolve. Therefore, studies like ours will need to be replicated and updated as longer-term longitudinal data become available. Second, ENDS use was only reported by a small number of respondents, potentially limiting the power to detect an association between ENDS use and COPD. PATH is a nationally representative sample of the US population, so ENDS use prevalence levels are reflective of the patterns of use in the population. However, ENDS use has increased recently, predominantly among youth and young adults,^35,36^ so further of PATH data as more waves become available might have larger samples of ENDS users. Third, the results from this study are based on self-reported incident diagnosed COPD. This is a limitation of the PATH data, but previous studies have shown high concordance between self-reported COPD and medical records.^37^

## Conclusions

Using nationally representative prospective data among US adults aged 40+, time-varying ENDS use during a five-year period did not increase the risk of self-reported incident diagnosed COPD once current cigarette use and cigarette pack-years were taken into account. Most adults who used ENDS either currently smoked cigarettes or smoked cigarettes in the past; this and the results from this study highlight the need to control for cigarette smoking history to assess any potential independent health effects of ENDS use on COPD.

## Take-Home Points

1. Study Question. Does electronic nicotine delivery systems (ENDS) use increase the risk of COPD independently of smoking history in a US nationally representative cohort?
2. Results. Current and former smoking and cigarette pack-years are strong determinants of COPD risk, but ENDS use, over a 5-year period, is not associated with COPD incidence after adjusting for smoking history.
3. Interpretation. It is critical to use prospective longitudinal data and properly control for cigarette smoking history to assess the independent health effects of ENDS.

## Data Availability

All data used is publicly available

## Supplemental Material

**Figure S1.**
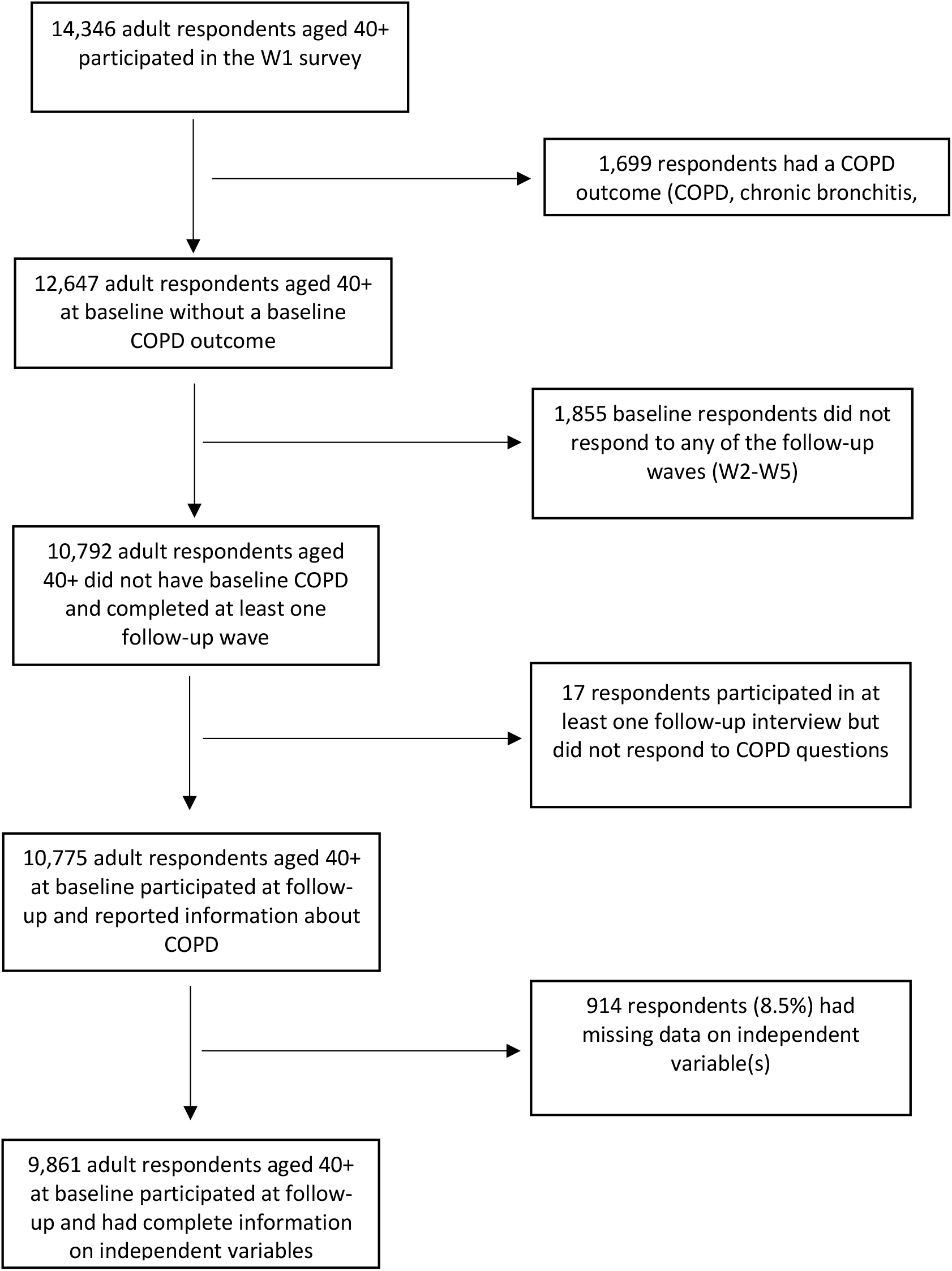
Flowchart of Sample Selection for Analytic Sample.

**Table S1.**
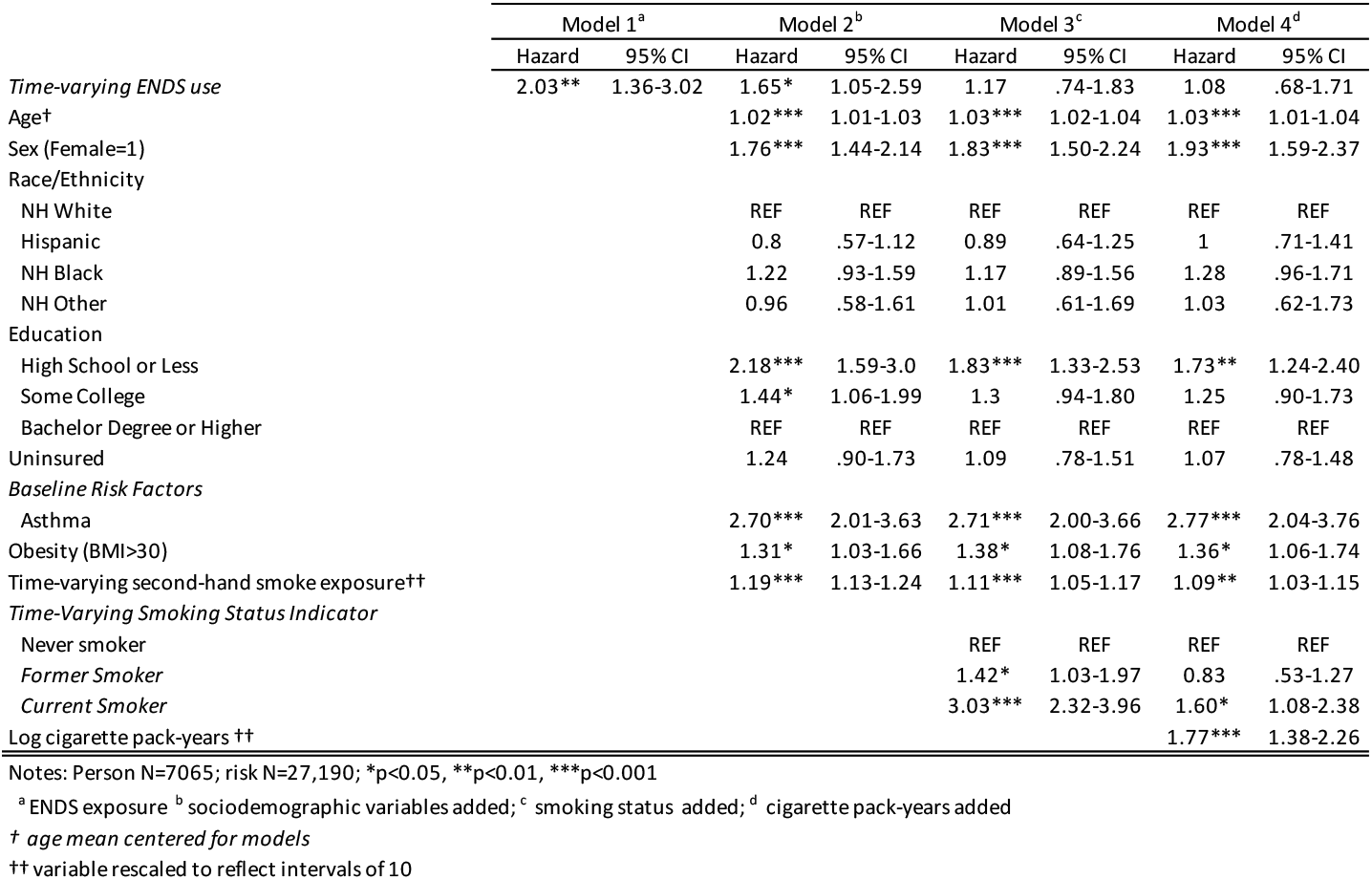
Discrete time survival analysis predicting incidence of self-reported chronic obstructive pulmonary disease (COPD) for longitudinal cohort (‘all waves’ weights) aged 40 and older, Population Assessment of Tobacco and Health Study (Waves 1-5, 2013-2019)

**Table S2.** Discrete time survival analysis predicting incidence of self-reported chronic obstructive pulmonary disease (COPD) for adults aged 40 and older who reported seeing a healthcare professional, Population Assessment of Tobacco and Health Study (Waves 1-5, 2013-2019)

|  | Model 1<sup>a</sup> |  | Model 2<sup>b</sup> |  | Model 3<sup>c</sup> |  | Model 4<sup>d</sup> |  |
| --- | --- | --- | --- | --- | --- | --- | --- | --- |
|  | Hazard | 95% CI | Hazard | 95% CI | Hazard | 95% CI | Hazard | 95% CI |
| *Time-varying ENDS use* | 1.96*** | 1.42-2.72 | 1.71** | 1.21-2.41 | 1.19 | 0.84-1.68 | 1.11 | 0.78-1.58 |
| Age† |  |  | 1.03*** | 1.02-1.04 | 1.03*** | 1.02-1.04 | 1.03*** | 1.02-1.04 |
| Sex (Female=1) |  |  | 1.61*** | 1.36-1.92 | 1.71*** | 1.43-2.04 | 1.82*** | 1.52-2.17 |
| Race/Ethnicity |  |  |  |  |  |  |  |  |
| NH White |  |  | REF | REF | REF | REF | REF | REF |
| Hispanic |  |  | 0.94 | 0.72-1.21 | 1.05 | 0.81-1.36 | 1.18 | 0.91-1.52 |
| NH Black |  |  | 1.19 | 0.96-1.47 | 1.14 | 0.92-1.42 | 1.25* | 1.0-1.56 |
| NH Other |  |  | 0.94 | 0.58-1.50 | 0.98 | 0.62-1.57 | 1 | 0.62-1.60 |
| Education |  |  |  |  |  |  |  |  |
| High School or Less |  |  | 2.17*** | 1.65-2.86 | 1.82*** | 1.39-2.39 | 1.70*** | 1.29-2.25 |
| Some College |  |  | 1.57** | 1.17-2.10 | 1.41* | 1.05-1.89 | 1.34 | 1.0-1.79 |
| Bachelor Degree or Higher |  |  | REF | REF | REF | REF | REF | REF |
| Uninsured |  |  | 1.30 | 0.96-1.75 | 1.16 | 0.86-1.56 | 1.15 | 0.85-1.54 |
| *Baseline Risk Factors* |  |  |  |  |  |  |  |  |
| Asthma |  |  | 2.95*** | 2.26-3.86 | 2.99*** | 2.29-3.91 | 3.05*** | 2.32-4.01 |
| Obesity (BMI>30) |  |  | 1.35*** | 1.11-1.63 | 1.43*** | 1.18-1.73 | 1.41** | 1.15-1.71 |
| Time-varying second-hand smoke exposure†† |  |  | 1.18*** | 1.14-1.23 | 1.11*** | 1.06-1.16 | 1.09*** | 1.04-1.14 |
| *Time-Varying Smoking Status Indicator* |  |  |  |  |  |  |  |  |
| Never smoker |  |  | REF | REF | REF | REF | REF | REF |
| Former Smoker |  |  |  |  | 1.50** | 1.17-1.94 | 0.85 | 0.49-1.22 |
| Current Smoker |  |  |  |  | 3.08*** | 2.51-3.78 | 1.58** | 1.12-2.23 |
| Log cigarette pack-years†† |  |  |  |  |  |  | 1.80*** | 1.47-2.21 |

Notes: Person N=9,875; risk N=33,752; \*p<0.05, \*\*p<0.01, \*\*\*p<0.001
<sup>a</sup> ENDS exposure <sup>b</sup> sociodemographic variables added; <sup>c</sup> smoking status added; <sup>d</sup> cigarette pack-years added
† age mean centered for models
†† variable rescaled to reflect intervals of 10

**Table S3.** Discrete time survival analysis predicting incidence of self-reported chronic obstructive pulmonary disease (COPD) for respondents aged 40 and older with regular ENDS use (10+ days per month), Population Assessment of Tobacco and Health Study (Waves 1-5, 2013-2019)

|  | Model 1<sup>a</sup> |  | Model 2<sup>b</sup> |  | Model 3<sup>c</sup> |  | Model 4<sup>d</sup> |  |
| --- | --- | --- | --- | --- | --- | --- | --- | --- |
|  | Hazard | 95% CI | Hazard | 95% CI | Hazard | 95% CI | Hazard | 95% CI |
| *Time-varying ENDS use* | 1.95** | 1.34-2.84 | 1.66* | 1.12-2.47 | 1.23 | .83-1.82 | 1.16 | .79-1.71 |
| Age† |  |  | 1.03*** | 1.02-1.03 | 1.03*** | 1.02-1.04 | 1.03*** | 1.02-1.04 |
| Sex (Female=1) |  |  | 1.60*** | 1.34-1.89 | 1.68*** | 1.41-2.01 | 1.79*** | 1.50-2.13 |
| Race/Ethnicity |  |  |  |  |  |  |  |  |
| NH White |  |  | REF | REF | REF | REF | REF | REF |
| Hispanic |  |  | 0.94 | .73-1.21 | 1.06 | .82-1.36 | 1.18 | .92-1.52 |
| NH Black |  |  | 1.21 | .99-1.49 | 1.17 | .94-1.44 | 1.28* | 1.03-1.58 |
| NH Other |  |  | 0.94 | .59-1.49 | 0.99 | .63-1.56 | 1.01 | .64-1.59 |
| Education |  |  |  |  |  |  |  |  |
| High School or Less |  |  | 2.20*** | 1.67-2.88 | 1.83*** | 1.40-2.40 | 1.72*** | 1.30-2.25 |
| Some College |  |  | 1.57** | 1.17-2.10 | 1.40* | 1.05-1.88 | 1.33* | 1.0-1.78 |
| Bachelor Degree or Higher |  |  | REF | REF | REF | REF | REF | REF |
| Uninsured |  |  | 1.32 | .99-1.77 | 1.18 | .88-1.57 | 1.16 | .88-1.55 |
| *Baseline Risk Factors* |  |  |  |  |  |  |  |  |
| Asthma |  |  | 2.94*** | 2.26-3.84 | 2.99*** | 2.29-3.90 | 3.05*** | 2.33-4.0 |
| Obesity (BMI>30) |  |  | 1.34** | 1.11-1.63 | 1.43*** | 1.18-1.73 | 1.41** | 1.16-1.71 |
| Time-varying second-hand smoke exposure†† |  |  | 1.19*** | 1.15-1.23 | 1.11*** | 1.06-1.16 | 1.09*** | 1.04-1.14 |
| *Time-Varying Smoking Status Indicator* |  |  |  |  |  |  |  |  |
| Never smoker |  |  |  |  | REF | REF | REF | REF |
| Former Smoker |  |  |  |  | 1.50** | 1.17-1.94 | 0.85 | .59-1.23 |
| Current Smoker |  |  |  |  | 3.16*** | 2.58-3.87 | 1.64** | 1.17-2.29 |
| Log cigarette pack-years †† |  |  |  |  |  |  | 1.79*** | 1.46-2.19 |

Notes: Person N=9,858; risk N=33,674; \*p<0.05, \*\*p<0.01, \*\*\*p<0.001
<sup>a</sup> ENDS exposure <sup>b</sup> sociodemographic variables added; <sup>c</sup> smoking status added; <sup>d</sup> cigarette pack-years added
† age mean centered for models
†† variable rescaled to reflect intervals of 10

